# Electronic data review, client reminders, and expanded clinic hours for improving cervical cancer screening rates after COVID-19 pandemic shutdowns: a multi-component quality improvement program

**DOI:** 10.1101/2023.01.20.23284607

**Authors:** Sue Ghosh, Jackie Fantes, Karin Leschly, Julio Mazul, Rebecca Perkins

## Abstract

**Objective:** To improve cervical cancer screening (CCS) rates, the East Boston Neighborhood Health Center (EBNHC) implemented a Quality Improvement (QI) initiative from March to August 2021.

**Methods:** Staff training was provided. A 21-provider team validated overdue CCS indicated by electronic medical record data. To improve screening, CCS-only sessions were created during regular clinic hours (n=5) and weekends/evenings (n=8). Patients were surveyed on their experience.

**Results:** 6126 charts were reviewed. Of the list of overdue patients, outreach was performed to 1375 patients to schedule the 13 sessions. A total of 459 (33%) of patients completed screening, 622 (45%) could not be reached, and 203 (15%) canceled or missed appointments. The proportion of total active patients who were up to date with CCS increased from 68% in March to 73% in August 2021. Survey results indicated high patient satisfaction, and only 42% of patients would have scheduled CCS without outreach.

**Conclusions:** The creation of a validated patient chart list and extra clinical sessions devoted entirely to CCS improved up-to-date CCS rates. However, high rates of unsuccessful outreach and cancelations limited sustainability. This information can be used by other community health centers to optimize clinical workflows for CCS.

**Funding:** All funding was internal from EBNHC Adult Medicine, Family Medicine, and Women’s Health Departments.

## INTRODUCTION

Nationwide closures at the beginning of the COVID-19 pandemic led to decreases in breast, colorectal, and cervical cancer screenings between 86% and 94% compared to three-year averages (Mast, 2022). The postponed screenings have led to backlogs that health care systems will need to address as operational changes continue with evolving COVID-19 rates. Federally Qualified Health centers (FQHCs), which primarily serve low-income and minority communities, have been particularly impacted by the pandemic. Low-income and minority communities had disproportionately high cancer incidence and mortality prior to the pandemic (Du, 2011); and now could have increased disparities due to reduced access to screening because of COVID-19 (Maringe, 2020).

East Boston Neighborhood Health Center (EBNHC) is the largest federally qualified health center (FQHC) in Massachusetts. It was established in 1970 and has approximately 170,000 patient visits annually. Its catchment area includes 270,000 patients. Over 70% of the patient population is Latinx. During the first year of the COVID-19 pandemic, most non-urgent in person medical care was postponed, including cervical cancer screening (CCS). A quality-improvement project was initiated at EBNHC in March 2021 to improve CCS rates. The multi-component intervention utilized best practices as recommended by the Center for Disease Control and Prevention’s (CDC) Community Preventive Services Task Force community guide including interventions to increase community demand (client reminders), interventions t increase community access (extended hours), and interventions to increase provider delivery of screening services (provider assessment and feedback) (CDC, 2019). This manuscript describes the project’s background, lessons learned, and new initiatives EBNHC will use to improve CCS rates in the future.

## METHODS

### Preparatory work: Demonstrating project need

An EPIC work-bench report (WBR) was run to identify active EBNHC patients (defined as at least 1 visit to a primary care department in the past 18 months) whose health record stated that CCS screening was overdue. This list had over 7000 patients as of January 2021. The provider tasked with examining EBNHC’s overdue CCS issue reviewed 1600 charts and noted that over 80% were correctly identified as overdue. These data were presented to EBNHC clinical and administrative leadership in February 2021. A proposal was made to 1) validate the overdue CCS list, 2) create CCS-only clinics, and 3) improve clinical and electronic CCS workflows. This project was approved by the Project Steering Committee March 2021. This manuscript reports efforts on chart validation and CCS-only clinics.

### Data Validation: Creation of outreach list

Starting in March 2021, a 21 provider team was created to review 6126 charts from the work-bench report generated by the Epic EMR system for overdue CCS. The team consisted of nurse practitioners, physician assistants, and physicians from Adult Medicine, Family Medicine, and OB/GYN departments. The team was trained on how to review charts to confirm need for CCS by the project lead during a one hour teaching session. One hundred medical record numbers from the overdue CCS list were sent approximately every two weeks to each provider to review. A new list of patients was sent to the providers once they reviewed the previously given list.

This was completed from April 30 – June 29, 2021, until the entirety of the overdue CCS list was reviewed. All providers received overtime pay from their respective departments for the hours used to review these patient records. A messaging pool was created in the electronic medical record (EMR) for providers to ask gynecologists questions about CCS necessity or frequency during the validation process. Three staff gynecologists volunteered for this task. A validated list was created of patients overdue for CCS.

### Staff training: Increasing awareness among clinicians

The importance of the problem, magnitude of the deficit, and goals of the QI project were presented to all staff during quarterly staff meetings by the Chief Medical Officer.

### Patient services: CCS-only Clinics

To address the backlog of patients overdue for CCS, screening-only clinics were created. To maximize accessibility for patients, two different time blocks were used: regular clinic hours and evenings/weekends.

#### Regular clinic hours

From March – April 2021, the OB/GYN department organized five CCS-only clinic sessions during regularly scheduled clinical hours. The OB/Gyn staff (medical assistants and front desk staff) outreached to patients from the validated list. Because these clinics occurred during regular clinic hours, the normal workflow for patient outreach and scheduling was used and no additional staffing or training was required. The patients outreached for these clinics all had a history of abnormal CCS.

#### Evenings/weekends

From May – July 2021, eight additional CCS-only clinic sessions were organized outside of regularly-scheduled clinical hours (evening/weekends). To organize and to run the evening/weekend CCS-only sessions, an operations team was created. The team consisted of the CCS project lead, the OB/Gyn clinical leader, operations managers from OB/Gyn and Family Medicine, and the OB/Gyn clinical supervisor. Weekly meetings were held from April 2 – July 23, 2021. A dedicated outreach team was also created from the OB/Gyn department: an outreach lead (OB/Gyn clinical supervisor), a medical assistant, and two front desk staff. The outreach team was responsible for calling patients from the validated list to schedule and to keep track of outreach outcomes. A smart phrase was created for these outreach calls. Staff also placed CCS clinic reminder calls one day before the scheduled CCS-only clinic. If a patient preferred to have her CCS done by her primary care provider, the message created by the new smart phrase was forwarded to the appropriate departmental pool to inform them to call the patient. The IT department created a new resource schedule in the Family Medicine department for these clinics. The data management and analysis were done by the outreach and project leads.

The staffing for the evening/weekend CCS-only clinics was organized by the project lead. Two rotating OB/Gyn front desk staff were used for front desk staff. Six rotating medical assistants and fourteen staff providers (NPs, PAs, and MDs) from OB/Gyn, Family Medicine (FM), and Adult Medicine (AM) departments were used. Three FM MD residents and two FM NP residents also participated. All clinic staff were paid overtime for the hours spent in the clinic (residents are not clinic staff and did not receive overtime pay). An orientation document regarding the CCS-only clinics was created by the project lead and emailed to participating providers several days before the clinic to review. A smart phrase for a CCS-only EMR note was also created for those providers who wanted to use it.

The evening/weekend sessions were done at different times to evaluate optimal timing and staffing for possible future permanent CCS clinics. All clinics were held in the Family Medicine clinic due to many exam rooms. A total of 4 Wednesday clinics were held from 5:20 – 8:00 pm; each included 48 appointments. Staff for Wednesday even clinics included six providers, four medical assistants, and one front desk staff. A total of 4 Saturday morning clinics were held from 8:00 am – 12:00 pm. Staff included five providers, 3 medical assistants, and one front desk staff. The first two Saturday clinics had 72 appointments. The last two Saturday clinics were overbooked with 73 and 79 patients respectively based on the number of missed appointments.

### Patient Satisfaction

The operations team wanted to collect patient satisfaction data on the evening/weekend CCS clinics to improve clinics in the future. A patient survey was created with leaders of the CCS project operations team, the director of clinical compliance and risk management and the Crossroads Group survey company. The survey was created in English and Spanish and was handed out to patients when they were being roomed. Following their visits, patients placed the completed surveys into a marked box, and the surveys were subsequently sent to The Crossroads Group for data analysis.

The Project Steering and Patient Care Committees at EBNHC gave ethical approval of this quality improvement project. Permission to report de-identified aggregate data and operational details were given by the Chief Medical and Chief Quality Officers.

## RESULTS

### Data validation/Reason for missed CCS

Because all active EBNHC patients had at least one visit in primary care the past 18 months, all had theoretically one or more opportunities for CCS. Therefore, 118 charts were randomly selected for a closer review to evaluate why CCS was not done. Among these 118 patients, 20% did not need a CCS: 14% due to incorrect CCS frequency in the health care gap; 3% of patients had CCS done at an outside clinic which was not seen in patient EBNHC chart; 3% of patients were not part of the EBNHC active patient panel; 1% of patients were > 65 years old or was status post a hysterectomy and did not have CCS health gap turned off. Among those who were due for CCS, several reasons were identified for not completing the screening. In 30% of encounters, the overdue CCS was not noted by the provider, while in 16% of encounters, the overdue CCS was noted but not addressed due to other medical concerns. Patient-related issues were noted in the remaining 54% of cases: not emotionally ready (23%), desired gynecologist (14%), on menses (9%), desired female provider (6%), and physical and cognitive impairments (2%).

### CCS results and rates

A total of 459 CCS’s were done during this project. This included 126 CCS during regular clinic sessions, 287 done during evening/weekend clinics, and 46 done by EBNHC PCPs during the project period. The results were as follows: 380 (83%) NILM HPV neg; 16 (3%) ASCUS HPV neg ; 20 (4%) NILM HPV +; 11 (2%) ASCUS HPV+; 5 (1%) LSIL HPV neg; 10 (2%) LSIL HPV +; 1 (<1%) HSIL HPV + HPV 16 neg; 10 (2%) Insufficient; 3 (<1%) not resulted; 3 (<1%) ordered but not done; 1 (<1%) not done (FIGURE 1).

**FIGURE 1:**
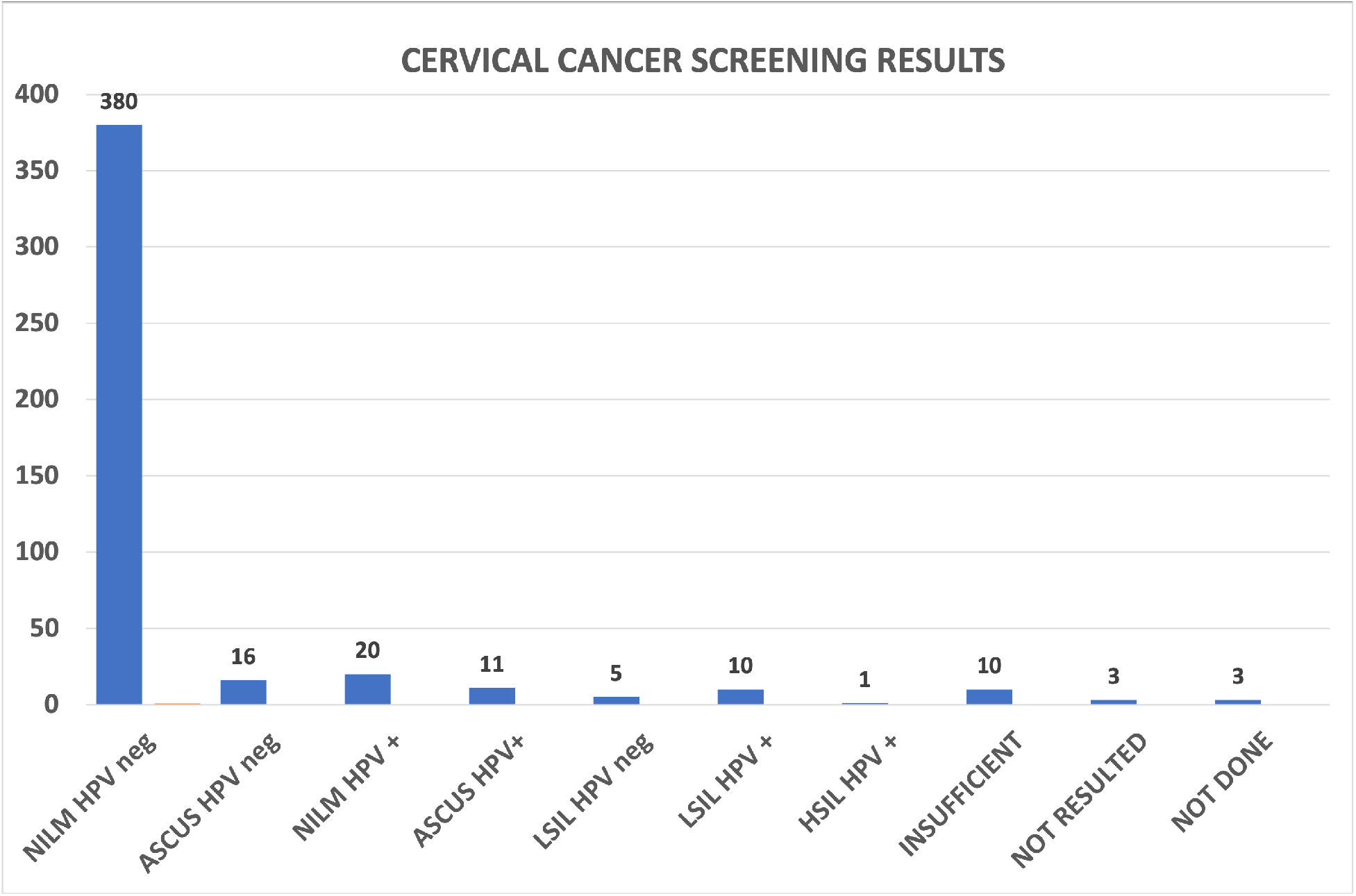
CCS RESULTS

The percentage of all active patients at EBNHC defined as having had a primary care visit in the past 18 months who were up to date with cervical cancer screening was 63.5% (36,824 active patients up to date on CCS /57,965 active patients who are eligible for CCS) in October 2020 (nadir of the pandemic), and 68.2% at the start of the project in March 2021 (36,432 /53,457).

Following the months of the data validation and CCS clinics, the up-to-date screening rate in August 2021 increased to 72.7% (39,040/53,678)(FIGURE 2).

**FIGURE 2:**
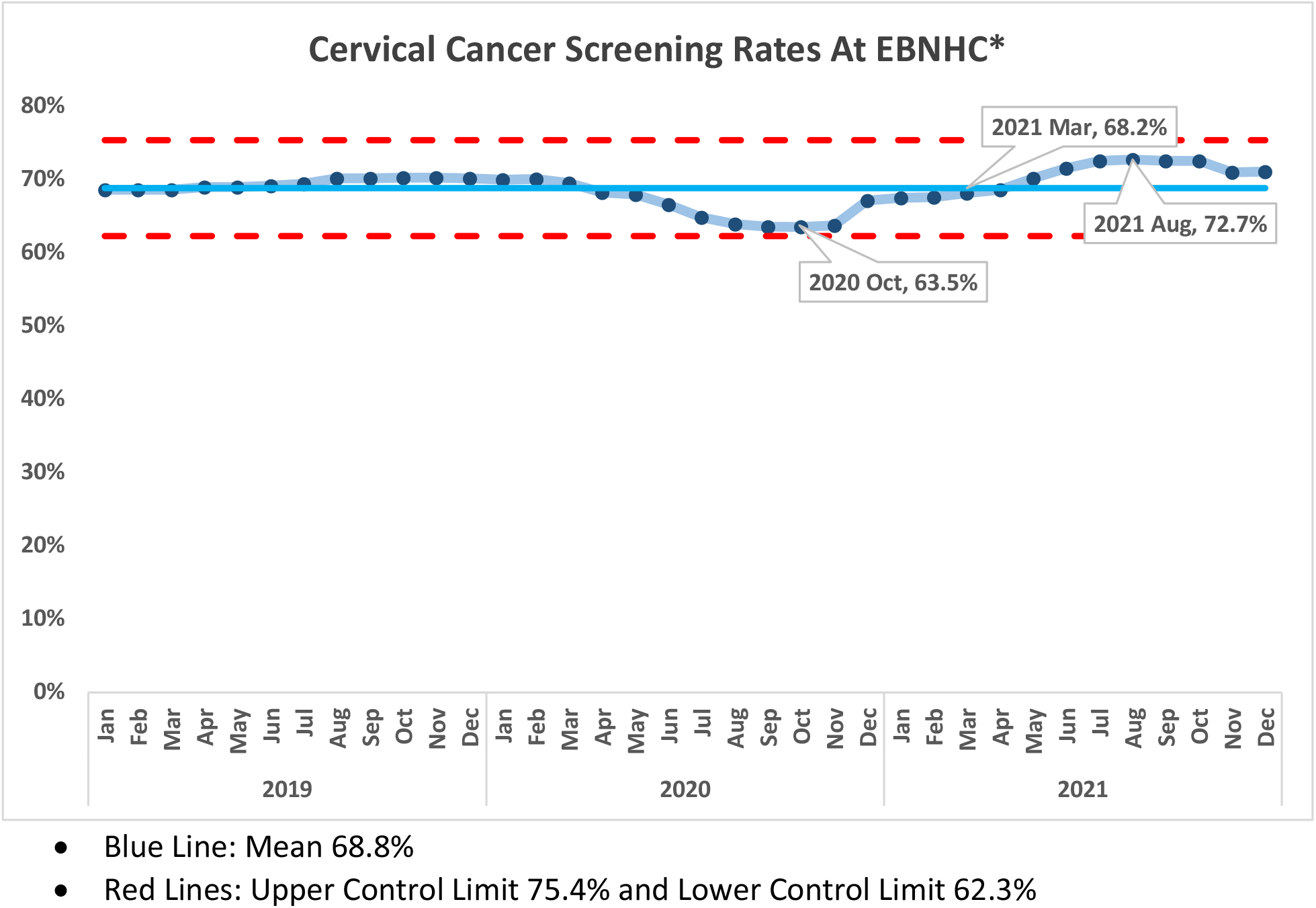
CCS RATES AT EBNHC

#### CCS-only Clinics

The CCS-only clinics were effective in addressing the backlog of patients. However, they involved a significant investment of clinic resources for staffing and outreach, and many patients could not be reached or missed their scheduled appointments.

#### Regular Clinic Hours

The newly created Patient outreach team initially called 220 patients all with a history of abnormal CCS. In total, 126 (57%) patients had CCS done. 49 CCS were done during CCS-only clinics in the Gynecology Department. The remaining 77 patients were seen by various Gynecology providers during their regularly scheduled clinics. 82% of patient attended scheduled appointments (126/153). Anecdotally, patients and providers felt the CCS clinics were done efficiently. Fifteen percent of patients wanted to discuss other gynecologic issues during their CCS visit which was accommodated. Among the 94 patients who did not complete CCS, 22% patients were not reached, and a letter was sent, or a voicemail was left, 27 (12%) canceled or missed their appointments, 3% declined screening, 3% had their CCS done at an outside clinic, 1% moved, and < 1% of patients wanted to have their CCS done by their PCP. (FIGURE 3)

**FIGURE 3:**
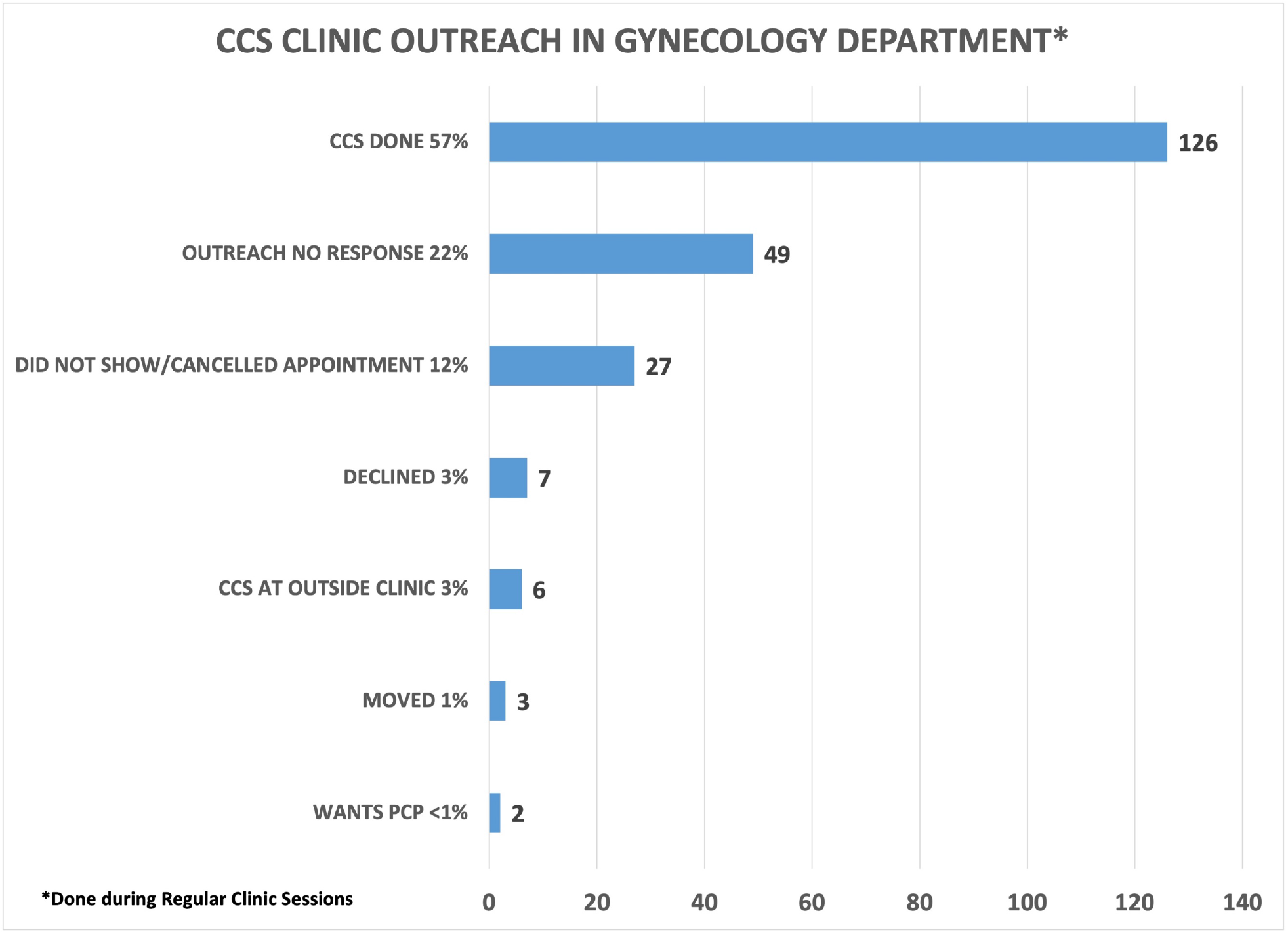
CCS CLINIC OUTREACH IN GYNECOLOGY DEPARTMENT

**FIGURE 4.**
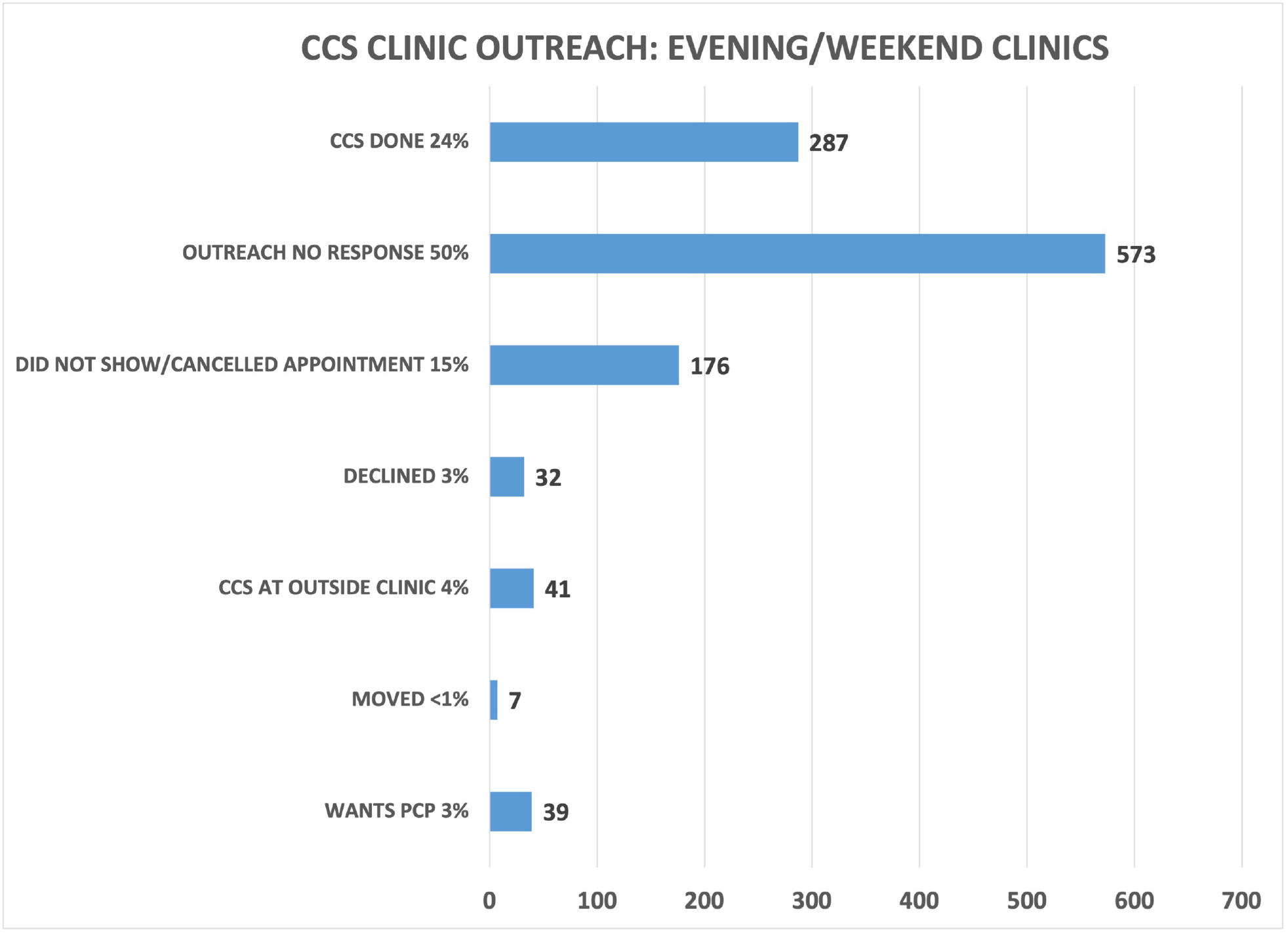
CCS CLINIC OUTREACH FOR EVENING/WEEKEND SESSIONS Footnote: For the patients who wanted their CCS to be done by their PCP, we created a workflow where the patient outreach team member could send a message in the EMR to the PCP department to notify them of the patient’s desire for a CCS appointment. The PCP clinical team would then reach out to the patient to schedule it. Review of this workflow revealed 34 messages sent to PCP departments. 25 (74%) CCS were completed by PCPs: 14 AM patients and 11 FM patients. For the 15 patients who did not have CCS done, in Adult Medicine 3 letters/call were done; 4 patients were not outreached, and 6 patients declined CCS. For Family Medicine, 4 patients did not have CCS done: 1 letter/call; 2 patients were not outreached, 1 patient declined. For Pediatrics, the 2 patients had both physical and mental disabilities which would make a CCS difficult and the ongoing discussion about CCS will continue.

#### Evening/Weekend

A total of 1155 patients were called from the validated overdue CCS list, which included patients with and without prior abnormal results who were overdue for CCS. These patients were scheduled into one of 8 extra clinical sessions. Of these, 24% of patients completed their CCS, 50% were not reached, 15% canceled or did not show up for their appointment, 4% had the CCS done at an outside clinic, 3% wanted their PCP to do their CCS, 3% declined screening, and <1% had moved.

Among the 462 patients with scheduled visits, approximately 62% completed screening and 38% canceled or did not show up to their appointments. We examined whether there was a pattern for the missed appointments at each time slot to decide whether modifying the number of patients per time slot would improve attendance. Examining the evening and Saturday morning missed appointments did not show a pattern. (FIGURE 5-7)

**FIGURE 5:**
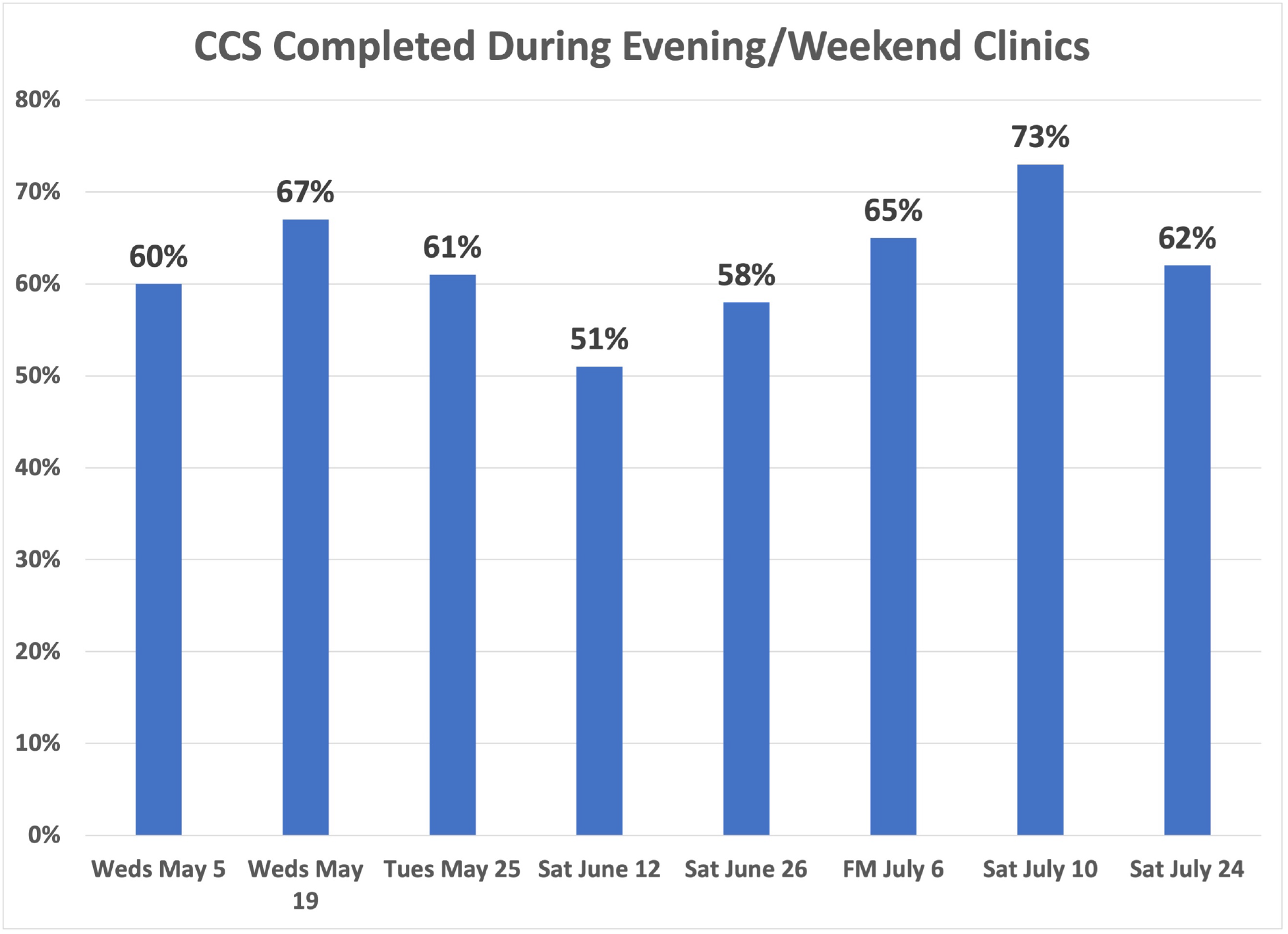
% CCS COMPLETED DURING EVENING/WEEKEND CLINICS* *CCS completion rates are reported among the patients who were scheduled into the evening/weekend clinics

**FIGURE 6:**
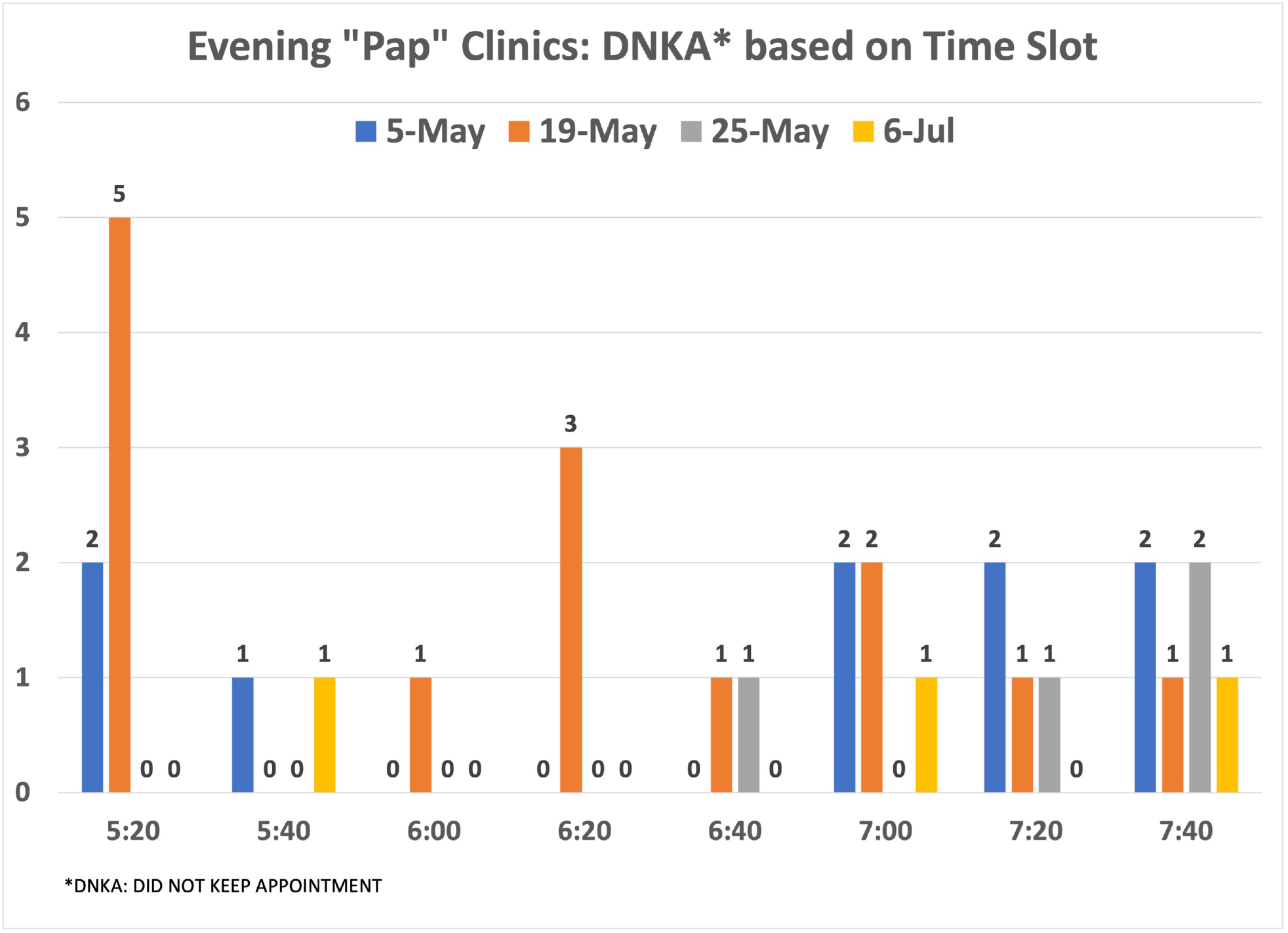
DISTRIBUTION OF NO-SHOW PATIENTS BASED ON TIME SLOT: EVENING CLINICS

**FIGURE 7:**
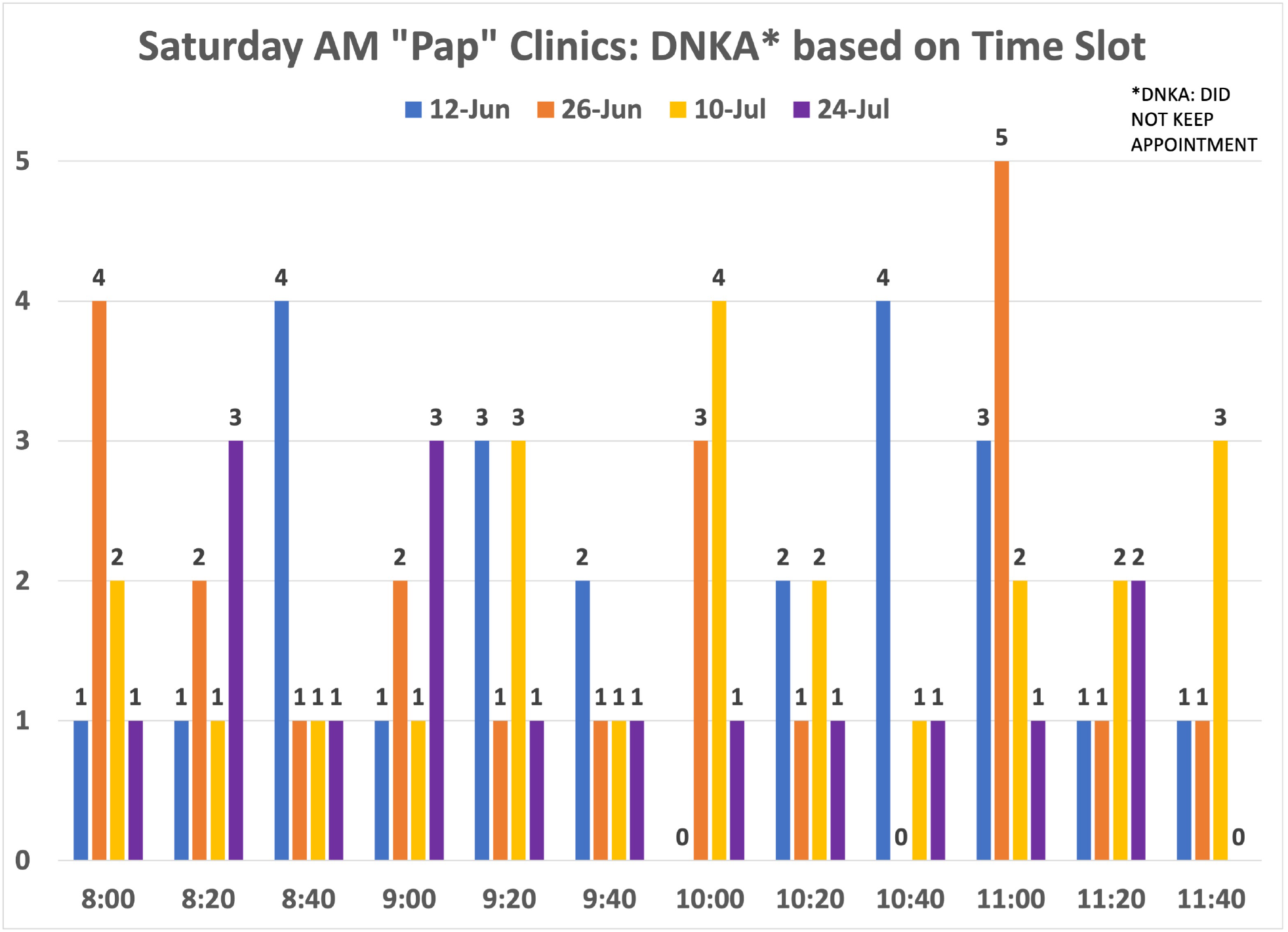
DISTRIBUTION OF NO-SHOW PATIENTS BASED ON TIME SLOT: SATURDAY AM CLINICS

### Patient Satisfaction

Patient satisfaction scores were 83% excellent (121/146 surveys) and 17% good (n=25/146 surveys). Patients largely rated their interactions as excellent with phone attendants (86%, 166/193) and healthcare providers (85%,162/191), and (77%, 146/190) found the test to be convenient. Nearly all, (93%, 168/181) reported that their expectations were met, and 95% stated they were very likely to make another CCS appointment in the future (95%,134/141). Only 42% (69/166) stated that they would have scheduled a CCS appointment without EBNHC outreach. Note that denominators are different as not every patient who turned in survey answered all the questions or the same questions.

Varying levels of work were needed to fulfill the objective of the project: improving CCS rates. A summary of this and future areas of work are in Table 1.

**TABLE 1:**
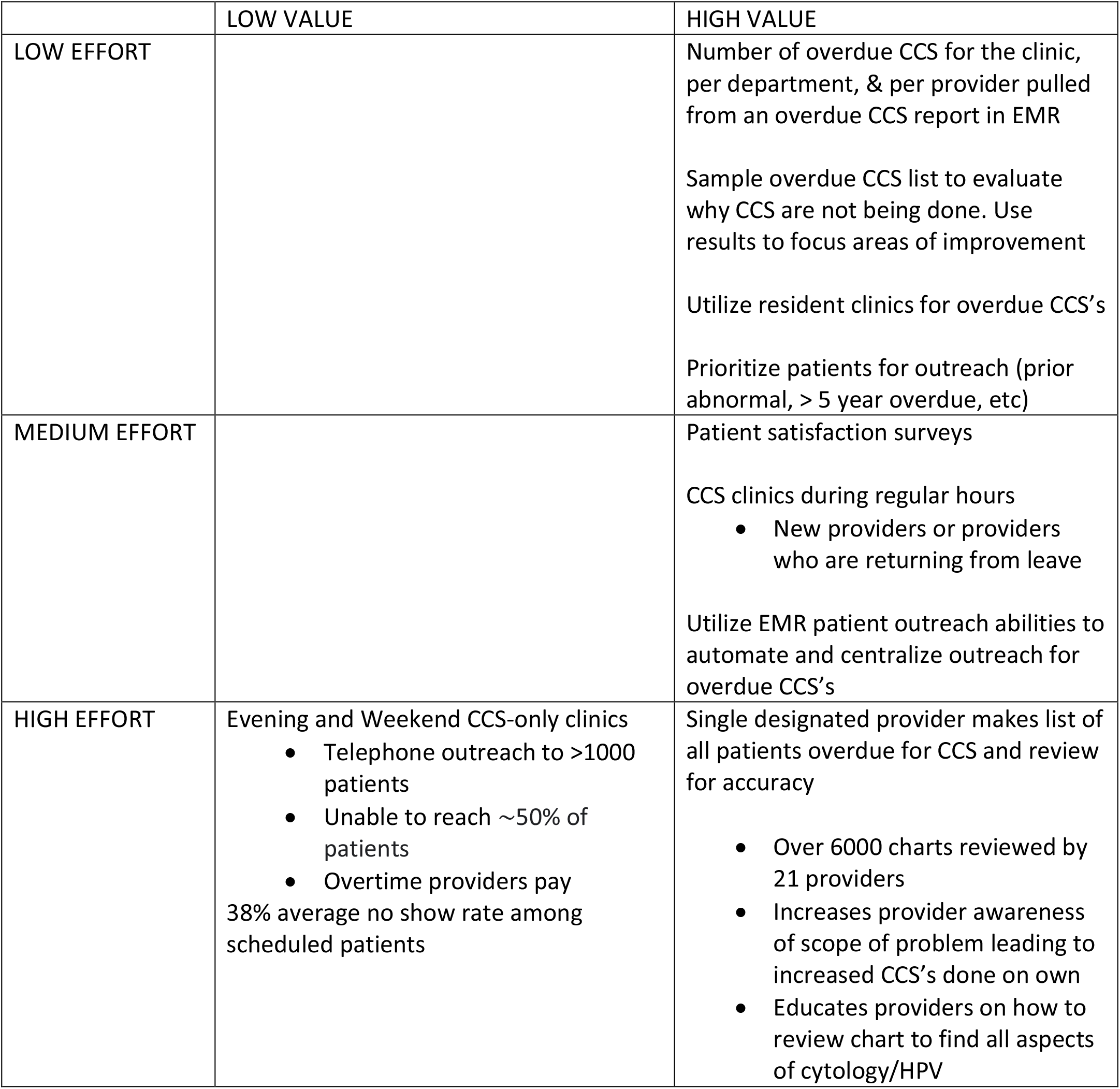
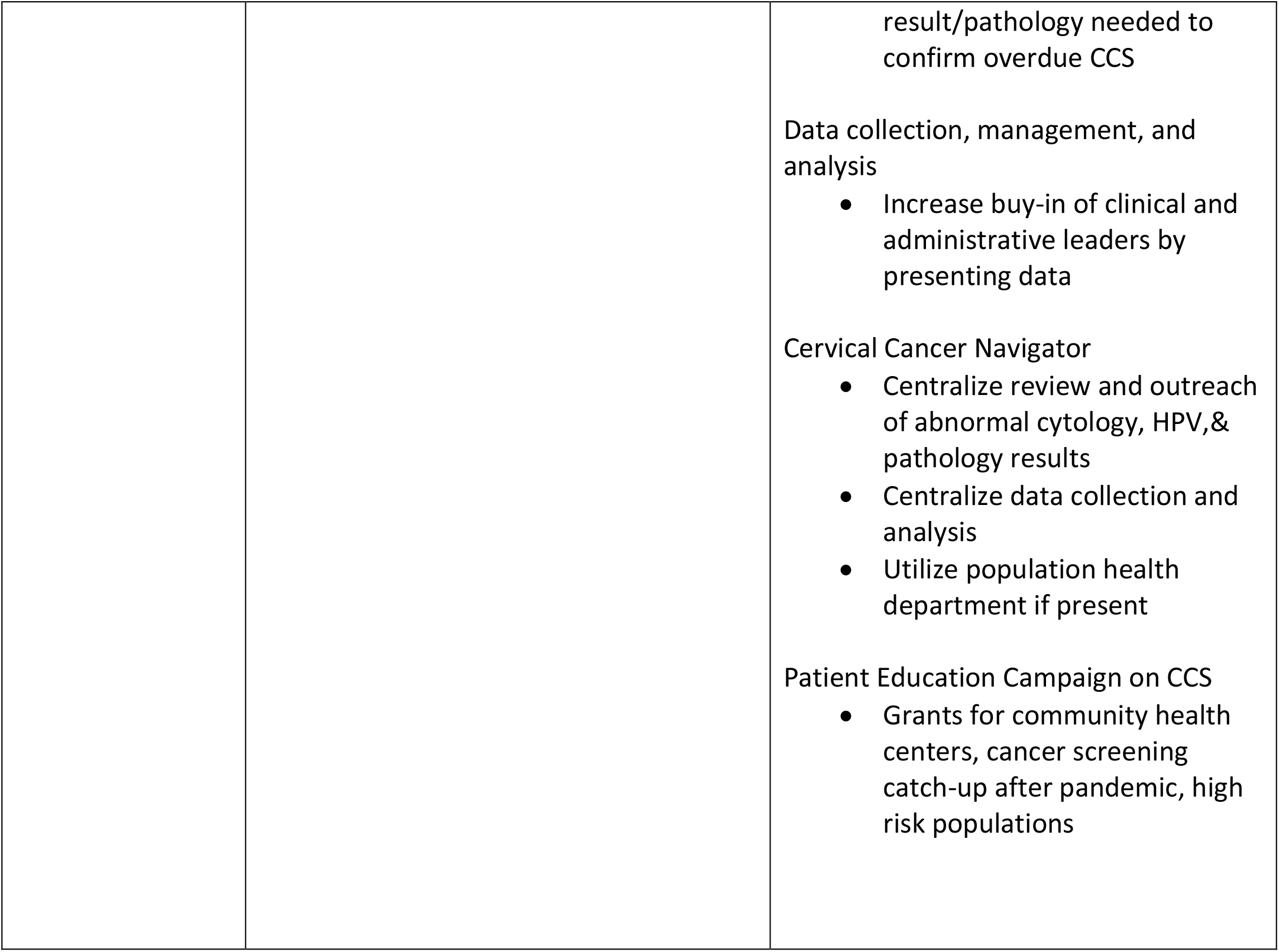
EFFORT VS VALUE SUMMARY FOR OVERDUE CCS PROJECT

**Table 2.** describes the lessons learned from this project that may be useful for other community health centers.

|  |
| --- |
| TABLE 2 LESSONS LEARNED |
| HOW MANY OVERDUE CCS'S DOES YOUR INSTITUTION HAVE? |
| - Examine details about the numbers: # Overdue CCS/provider and # Overdue CCS/department to see if education is needed regarding screening guidelines or clinical workflows |
| - Identify high risk groups to target patient outreach: no CCS for > 5 years, history of abnormal CCS, first CCS overdue |
| WHY IS CCS NOT BEING DONE? |
| - Random sampling of overdue list and deep dive into chart and identify why the CCS's were not done and change workflows accordingly. Changes in workflow should consider CCS-only clinics for new providers or providers coming back from leave. Evening/weekend CCS clinics are not necessarily the best use of resources. |
| IF YOUR CLINIC HAS A HIGH NO SHOW/CANCELLATION RATE FOR CCS, WHY? |
| - Electronic outreach and education, language specific messaging to these patients |
| - Survey these patients to see why they are not coming in for CCS |
| - Create a targeted Cervical Cancer Awareness campaign if possible |
| EDUCATE CLINICAL AND ADMINISTRATIVE LEADERS |
| - Use data from your clinic's overdue CCS list to obtain buy-in from strategic stakeholders who can support changes in clinical and electronic workflows |
| STANDARDIZE WORKFLOWS FOR RESCHEDULING PATIENTS WHO DECLINE/PROVIDER WHO CAN'T GET TO CCS DURING CLINIC VISIT |
| HAVE A CERVICAL CANCER NAVIGATOR/POPULATION HEALTH MANAGER TO OVERSEE ELECTRONIC OUTREACH AND DATA COLLECTION/ANALYSIS |

## DISCUSSION

EBNHC’s ability to increase the proportion of our active patient population up to date with CCS by 4.5% during the COVID-19 pandemic was multi-factorial. Validating overdue patient lists removed 15% of patients inaccurately flagged as needing screening, which created an accurate denominator to determine up-to-date status. Creating directed outreach and implementing CCS-only sessions led to completion of 459 CCS. In addition, return of patients to primary care as preventive services re-opened in after widespread COVID-19 vaccination in 2021; increased awareness among PCPs of the importance of CCS due to both direct staff outreach and participation in the chart validation; and support from clinical leadership may have led clinicians to capitalize on opportunities to perform CCS when patients presented for clinical care.

The CDC Community Preventive Services Task Force Community Guide recommends multicomponent interventions to increase cancer screenings (CDC, 2022). The main components of our intervention focused on increasing community demand (client reminders), interventions to increase community access (extended hours), and interventions to increase provider delivery of screening services (provider assessment and feedback). Establishing the intervention had several steps. First, we had to establish buy-in from leadership. Our patient population is > 70% Latinx. The Latina population have the highest incidence of new cervical cancer cases and the second (to black women) highest incidence of cervical cancer deaths in the US (CDC, 2019). We therefore felt this issue was a priority for our patient population. A single provider’s review of the overdue CCS list showed high rates of overdue screenings, which led to the buy-in from the clinical and academic leaders to proceed with the project. Understanding the scope of the issue and catching up with the CCS rate was important to prevent an increase in cervical precancer and cancer in our community.

Second, we had to determine which patients needed screening. There was an estimated 15% of the overdue CCS list which was incorrect based on random chart sampling. Our chart review of over 6000 charts created a validated list to use for focused electronic outreach in the future. It also created a greater understanding for providers of how to confirm and overdue CCS and to increase CCS during their clinics. Third, we had to raise awareness of the problem and of the goals of the QI project among clinic staff. Fourth, we had to develop efficient processes for completing overdue CCS. The evidence-based components chosen from the Community Guide included interventions to increase community demand (client reminders) and interventions to increase community access (extended hours) (CDC, 2019). After establishing the validated list and presenting the project at staff meetings (provider assessment and feedback), we provided client reminders (outreach) to patients as part of normal clinic workflows or via a newly created outreach team. To increase access (extended hours), we tried the following: 1) creating new CCS clinics during regular clinic hours in the Gynecology Department; 2) creating new CCS clinics during evening/weekend hours; 3) scheduling CCS during regular clinics in Gynecology Department. This included patients’ first scheduled appointments and rescheduled visits following missed appointments for the new CCS clinics or if patients called to schedule CCS after CCS clinics were finished. We also created workflows to communicate with Adult Medicine and Family Medicine Departments when patients preferred CCS to be done by the primary care provider.

We found that CCS-only clinics that focused on patients with prior abnormal results and were performed during regularly scheduled clinical hours were effective. The majority (57%) of patients who received outreach from the Gynecology department for CCS-only clinics during regular clinic hours had their screenings done. The success of these clinics may have occurred in part because all these patients had a history of abnormal CCS and may therefore have been more knowledgeable regarding the purpose and importance of CCS. These clinics were an effective way both to increase the CCS rate and to have new providers and providers coming back from leave ease into clinical work. Resident clinics were another way to improve CCS rates while teaching pelvic exams and being financially prudent. Surveying patients was a valuable tool for obtaining patient opinions for use in improving CCS clinics and workflow. Importantly, we found that fewer than half of patients would have scheduled CCS without outreach, underscoring the importance of increasing community demand, especially in safety-net settings.

There were aspects of this project which were less effective. Most surprising was that our attempts to increase access by offering CCS appointments during evening/weekend were not successful. Evening/weekend appointments had a much lower attendance rate (62%) compared with appointments during normal clinic hours (82%). Not all patients outreached for evening/weekend clinics had a history of abnormal CCS, therefore some may have been less knowledgeable regarding the importance of screening. Additional reasons for low attendance may have been lack of a direct recommendation for screening from their healthcare provider, fear of pain, and low perceived need especially during a pandemic. Ogebyte *et al* showed that a nurse contacting 120 patients overdue for CCS in a small practice in northwest England increased CCS rates versus texting, but the effort required to achieve this increase was unsustainable (Ogebyte, 2021). Another study of 260 patients showed a 8% increase in CCS for pregnant and postpartum patients by introducing a package of CCS information, targeted education, and widening access to screening appointments (Coleridge, 2022). Other research on CCS during the COVID-19 pandemic used different changes in workflow to improve screening rates. Martellucci et al. changed CCS appointment times from flexible scheduling for many patients in one time slot to strict 15 minute appointments for one patient only. This led to similar screening rate to pre-COVID and higher provider satisfaction (Martellucci, 2021). Castanon *et al*. demonstrated modeling recovery strategies for CCS emphasizing increased access and patient messaging (Castanon, 2021).

Future efforts to improve CCS at EBNHC will include automated electronic targeted outreach to specific patient groups (e.g., initial screening, screened > 5 years ago, and history of abnormal CCS). Optimizing the utilization of the features of an EMR system can reduce the need for work hours needed and improve the efficiency of data management and analysis. Instead of a single provider reviewing the overdue CCS EMR list, having a population health manager or grant-sponsored volunteer do the data management and analysis as a Cervical Cancer Navigator can be more efficient. Care navigation has been shown to increase cancer screening rates (Nelson, 2020). We also want to survey patients who would not schedule CCS or missed CCS appointments to create a community-specific Cervical Cancer Screening Campaign, including evaluating the role of social determinants of health. To reduce the number of patients requiring CCS each year, EBNHC has also recently updated their CCS screening guidelines to every three years for 21-24 year-olds with cytology only and every 5 years for 25-65 year-olds with HPV/cytology co-testing, consistent with American Cancer Society’s 2020 guidelines (Fontham, 2020). This will allow the extension of screening intervals from 3 to 5 years for most patients.

Eventual self-screening HPV testing could increase rates while optimizing resource allocation. We are currently not continuing CCS-only clinics due to the limited appointment access in all adult departments stemming from the increased need for in-person visits since the improvement of the COVID-19 pandemic. Restarting these clinics during regular hours in Family Medicine, Adult Medicine, and the OB/GYN departments is a future aim.

This study has both strengths and limitations. Our experience working from a list of over 7000 patients is larger than similar QI projects reported in the literature. This is also one of the first successful QI projects to our knowledge specifically addressing COVID-related screening deficits in a safety net setting. However, as we describe the experience in one FQHC, our results may not be generalizable to other settings such as rural clinics or those without EMR capabilities. EBNHC has had a provider who has been able to lead the project from the initial review of the overdue CCS EMR list to organizing the CCS clinic staffing and to create new clinical and electronic workflows to use in the future. Having dedicated staff to manage CCS and cancer screening may not be feasible for many community health centers.

## CONCLUSION

During the project March – August 2021, EBNHC performed 459 CCS and increased the proportion of our total patient population who were up to date with screening by 4.5% from its nadir during 2021. The information gathered from our overdue CCS list was utilized to launch a multidisciplinary effort to learn why CCS was not being done and to validate our overdue numbers. We have increased the awareness of our overdue CCS issue and regarding the EMR review needed to confirm and overdue CCS in three departments. The screenings done during the CCS project plus increased provider awareness have contributed to our increased CCS rate. We are also in the process of centralizing our CCS workflow to decrease charting errors and make patient outreach more automated and efficient. If CCS-only clinics can be done during regular hours or resident clinics, they have value. The lessons learned from our effort can be used by other community health centers to improve CCS rates and decrease health inequities for high-risk populations in the US.

## Data Availability

All data produced has been submitted with the manuscript as supplemental information.

